# Serum Glycobiomarkers Defining Therapeutic Response to Intravenous Immunoglobulin in Chronic Inflammatory Demyelinating Polyneuropathy

**DOI:** 10.1101/2024.05.02.24306789

**Authors:** Soma Furukawa, Yuki Fukami, Hisatoshi Hanamatsu, Ikuko Yokota, Jun-ichi Furukawa, Masaya Hane, Ken Kitajima, Chihiro Sato, Keita Hiraga, Yuki Satake, Satoru Yagi, Haruki Koike, Masahisa Katsuno

## Abstract

**Background:** Glycosylation plays a crucial role in various pathologic conditions, including inflammation. This study conducted a comprehensive glycan analysis of serum to determine how glycan biomarkers are associated with the pathophysiology of chronic inflammatory demyelinating polyneuropathy (CIDP) and the effects of its treatment.

**Methods:** We comparatively analyzed *N*- and *O*-glycans in the pretreatment serum of 27 treatment-naïve patients with typical CIDP and age- and sex-matched 20 healthy controls (HC) using mass spectrometry. We determined the association between clinical parameters and glycans. Treatment response was defined according to the degree of improvement in the modified Rankin Scale 2 weeks after the first dose of intravenous immunoglobulin (IVIg), and the serum glycan and neurofilament light chain (NfL) levels were assessed at the baseline.

**Results:** Compared with the HC, the CIDP group demonstrated significantly lower levels of serum total *N*-glycans (CIDP, median 973.3 [IQR 836.2–1131.3] pmol/µL; HC, 1125.0 [1005.0–1236.2] pmol/µL; *p* < 0.05), especially sialylated *N*-glycans (CIDP, 898.0 [752.2–1037.2] pmol/µL; HC, 1064.4 [942.7–1189.8] pmol/µL; *p* < 0.01). In contrast, the *O*-glycan levels did not differ significantly between the two groups. Treatment response was associated with low *N*-glycan levels but not with the serum NfL levels. For individual glycans, low levels of Hex_2_HexNAc_2_NeuAc_2_ [α2,6/α2,6] + Man_3_GlcNAc_2_, α2,6-linked sialylated *N*-glycans, showed the treatment response group to have an area under the curve of 0.802 (p < 0.05).

**Conclusions:** Low levels of sialylated *N*-glycans may serve as a novel biomarker reflecting pathophysiology and therapeutic resistance in typical CIDP.

**KEY MESSAGE:**

- What is already known on this topic

Abnormal conformational changes in glycans of serum proteins are associated with the pathogenesis of inflammatory diseases. In a demyelinating mouse model, *N*-glycans suppress the activity of inflammatory helper T- and B-cells. A decrease in sialylated *N*-glycans of serum IgG-Fc in the serum of patients with CIDP correlates with disease severity, suggesting the potential of serum glycans as biomarkers for CIDP.

- What this study adds

In the patients with typical CIDP, serum total *N*-glycans, especially sialylated types, were significantly decreased, indicating a reduction in sialylated *N*-glycans derived from glycoproteins in CIDP. Moreover, lower levels of total *N*-glycans, particularly α2,6-sialylated *N*-glycans, were associated with reduced responsiveness to initial IVIg treatment.

- How this study might affect research, practice or policy

The study’s findings provide a new approach to exploring the immunological and therapeutic aspects of the role of glycans in CIDP. The decrease in serum total *N*-glycans, specifically sialylated types, may reflect an inflammatory pathophysiology in CIDP. Furthermore, it is suggested that these changes may serve as novel biomarkers to predict response to initial IVIg treatment.

## INTRODUCTION

Chronic inflammatory demyelinating polyneuropathy (CIDP) is the most common immune-provoked chronic inflammatory neuropathologic condition. Typically, it manifests with proximal and distal symmetric muscle weakness and a disease progression that lasts >8 weeks.^1^ Its pathophysiology is heterogeneous, and there is no gold standard for its diagnosis.^2^ Furthermore, the treatment schemes include immunomodulation with intravenous immunoglobulin (IVIg), corticosteroids, and plasma exchange, but despite their transient benefit, certain population of patients develop progressive axonal damage.^3^ Therefore, the objective and quantitative biomarkers that reflect the pathophysiology and treatment response of CIDP should be determined.^4, 5^

The blood biomarkers for CIDP are limited to a few proteins including plasma total tau and serum peripherin, none of which have been established as either diagnostic or prognostic predictors of the disease or for monitoring the treatment response.^6, 7^ Among the reported blood biomarker candidates, the neurofilament light chain (NfL) has been reported to be associated with axonal damage, but the results have not been completely validated because of the diversity of pathophysiology and treatment courses in patients with CIDP.^8^ Glycosylation is a common post-translational modification of proteins that occurs in the endoplasmic reticulum and Golgi apparatus.^9^ Abnormal conformational changes of *N*- and *O*-glycans of serum proteins have been implicated in the development of cancer and inflammatory diseases.^10–13^ Glycosylation modulates immune responses by regulating the development of thymocytes and differentiation of T-helper cells.^14^

In the nervous system, numerous glycoproteins have been detected in a myelin sheath. They are an integral part of myelin formation, maintenance and degeneration.^15, 16^ During the process of demyelination, the dysfunction of mitochondria occurs, leading to aggravated oxidative stress.^17, 18^ Consequently, a shift from aerobic to anaerobic metabolism occurs, depleting the hexosamine pathway. As a result, the biosynthesis of uridine diphosphate-GlcNAc, the precursor of *N*-glycans branching, is hindered. This likely underlies the decreased levels of total *N*-glycans during demyelination in the central nervous system, whereas such changes are unknown in the peripheral nervous system.^19^ The reduced sialylation of serum IgG-Fc has been reported in patients with CIDP,^20^ but glycosylation alteration of the entire protein structures in CIDP is yet to be confirmed.

We hypothesized that patients with CIDP have a wide range of glycan changes not only in IgG but also in other serum proteins that could be measured as biomarkers. Our research aimed to conduct a comprehensive glycoanalysis of sera from patients with CIDP and investigate novel glycan biomarkers that can define the pathophysiology of the condition and efficacy of its treatment.

## METHODS

### Participants

This study included consecutive patients with confirmed clinical diagnoses of CIDP at the Nagoya University Hospital between July 2012 and August 2021 in accordance with the European Academy of Neurology/Peripheral Nerve Society guidelines 2021.^21^ Most patients were directed from other hospitals for further diagnostics and therapy, and only those with typical CIDP were enrolled. We measured the antibodies against paranodal cell adhesion molecules (i.e., contactin-1 [CNTN1], neurofascin-155 [NF155], and contactin-associated protein1 [Caspr1]) in all patient sera using enzyme-linked immunosorbent assays in our laboratory.^22^ Patients with CIDP variants or autoimmune nodopathies expressing positive anti-CNTN1, NF155, or Caspr1 antibodies were excluded from the study. Furthermore, patients who had been subjected to therapy with immunomodulatory agents (i.e., IVIg, plasma exchange, corticosteroids, or immunosuppressive drugs) before baseline were excluded. The age- and sex-matched HC comprised the comparison group. The inclusion criteria for the control group covered no brain injuries and neurological diseases in medical history.

The study was performed in accordance with the Declaration of Helsinki and Ethical Guidelines for Medical and Health Research Involving Human Subjects endorsed by the Japanese government, with the approval from the Ethics Review Committee of Nagoya University Graduate School of Medicine. Before participating in the research, written informed consent of the whole sample was obtained after receiving the content information endorsed by the Ethics Review Committee, including the unfavorable and favorable aspects of the study and other relevant data.

### Sample collection and serum NfL analysis

Serum was collected from patients on admission or attendance at the Nagoya University Hospital. The blood tubes were promptly removed to the laboratory department of the hospital and subjected to centrifugation within 1 hour following collection. The serum was frozen and preserved at −80°C until use. The serum NfL protein concentrations were measured in duplicate using the Simoa HD-1 Analyzer (Quanterix, Lexington, MA, USA) and ultrasensitive paramagnetic bead-based enzyme-linked immunosorbent assay as performed by researchers blinded to all clinical data.

### Extraction, purification, and analysis of serum *N*- and *O*-glycans

The releasing of *N*-glycan from serum proteins was performed according to previous reports.^23^ Released *N*-glycans were purified by glycoblotting procedures combining with sialic acid linkage specific alkylamidation (SALSA).^24, 25^ After removal of excess reagents, *N*-glycans labeled with aoWR were subjected to matrix-assisted laser desorption-ionization time of flight mass spectrometry (MALDI-TOF MS) analysis. In *O*-glycomic analysis, the sialic acid linkage-specific derivatization was performed according to the previous report.^25^ Briefly, secretor proteins in serum (10 μL) were derivatized in a linkage-specific manner using the SialoCapper-ID Kit. Excess SALSA reagents were removed from SALSA-derivatized serum proteins by acetonitrile presipitation. *O*-Glycans were prepared by BEP method with minor modifications.^26^ Derivatized *O*-glycans were subjected to MALDI-TOF MS analysis.

### Clinical evaluations

We evaluated the muscle strength using the Medical Research Council (MRC) scale and disease severity using the modified Rankin Scale (mRS). Treatment response was outlined based on the improvement in mRS 2 weeks after the initial administration of IVIg (400 mg/kg/day for 5 days) as compared to baseline: the group with no improvement in the mRS score was categorized into the non-responder group, and that with an improvement of 1 or more was categorized into the responder group.

The patients underwent nerve conduction studies on four motor nerves (median, ulnar, tibial, and peroneal nerves) and three sensory nerves (median, ulnar, and sural) with surface stimulation and recording electrodes according to standard protocols. The parameters analyzed for the motor nerves included distal latency, compound muscle action potential (CMAP) amplitude, and conduction velocity, whereas those for the sensory nerves covered sensory nerve action potential (SNAP) amplitude and conduction velocity. The CMAP and SNAP amplitudes were evaluated from baseline to the first negative peak.

### Statistical analysis

The *m*/*z* values were used to generate the compositions of glycans detected by tandem MALDI-TOF MS and create spectra. The descriptive statistics was expressed as median (interquartile range [IQR]) for non-parametric continuous data. The intergroup comparisons of the latter values were performed using the Mann–Whitney U test. The differences in categorical variables were analyzed using χ² test. The receiver operating characteristic (ROC) curves were evaluated for the patients with CIDP and HC along with the non-responder and responder groups, and the area under the curve (AUC) values were computed. Multiple comparisons were conducted using analysis of variance with the Kruskal–Wallis test, and if a significant difference was observed, pairwise tests were performed between the two groups, with the significant probabilities corrected by Bonferroni adjustment. The Spearman rank correlation coefficient was used to estimate the correlation between parameters. *p*-values of < 0.05 and correlation coefficients (r) > 0.3 were regarded as statistically significant. Orthogonal partial least squares discriminant analysis (OPLS-DA) was conducted to assess the differences between the non-responder and responder groups using MetaboAnalyst 5.0. The variable importance in projection (VIP) values indicated the importance of glycans in the OPLS-DA model. Statistical calculations were performed using the Statistical Package for the Social Sciences V.29.0J software (IBM Japan, Tokyo, Japan).

## RESULTS

### Baseline characteristics of the participants

Figure 1 reveals the flow diagram of the enrolled study participants. Of the 81 consecutive patients with CIDP, we excluded 9 with autoimmune nodopathies, 33 who had been administered with immunomodulatory agents prior to baseline, and 12 with CIDP variants. Eventually, 27 treatment-naïve cases of typical CIDP and 20 age- and sex-matched HC were included.

**Figure 1.**
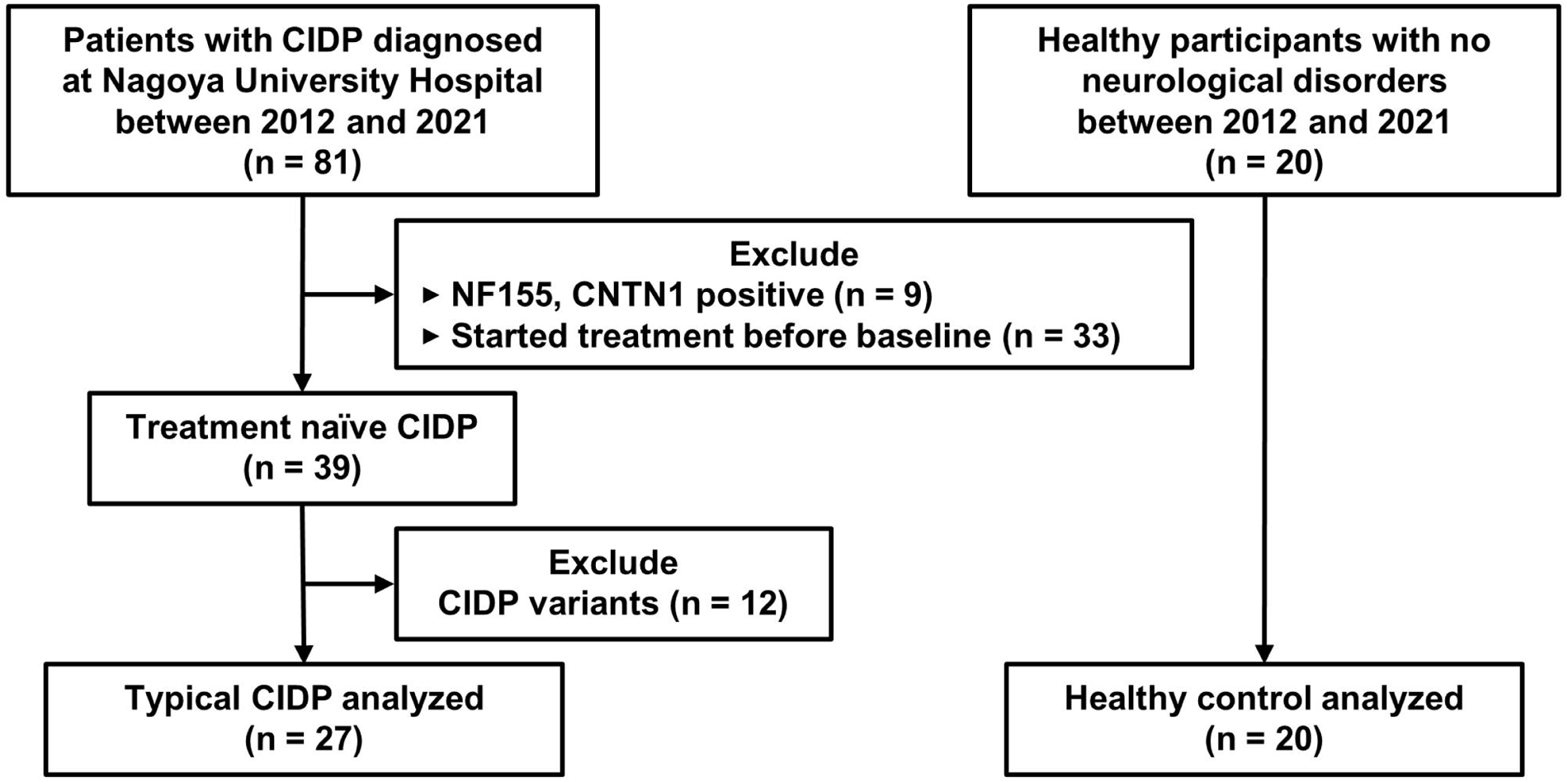
Flow diagram of the study subject enrollment. Enrollment flow diagram for patients with CIDP and HC. CIDP, chronic inflammatory demyelinating polyneuropathy. NF155, anti-neurofascin155 antibody; CNTN1, anti-contactin1 antibody.

Table 1 shows the clinical backgrounds of patients with CIDP and HC. The median mRS for patients with CIDP at baseline was 3. The patients with CIDP had significantly higher serum NfL levels than the HC (*p* < 0.05) and received initial treatment with IVIg. The degree of improvement in mRS at 2 weeks after the initial therapy as compared with the baseline values were 0 in 9 cases (33%), 1 in 12 cases (45%), and 2 in 6 cases (22%). No patient showed an improvement of ≥3.

**Table 1.**
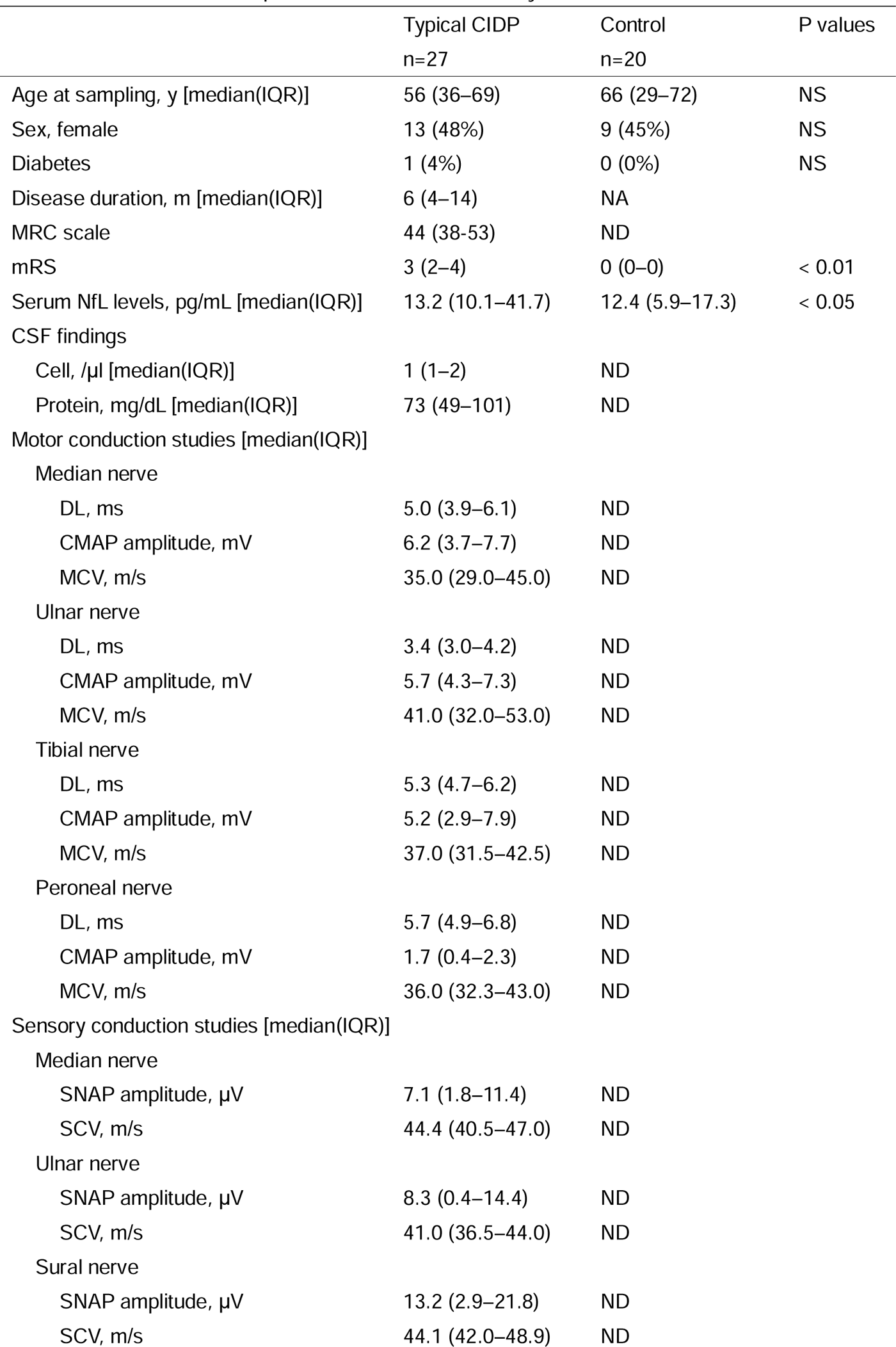
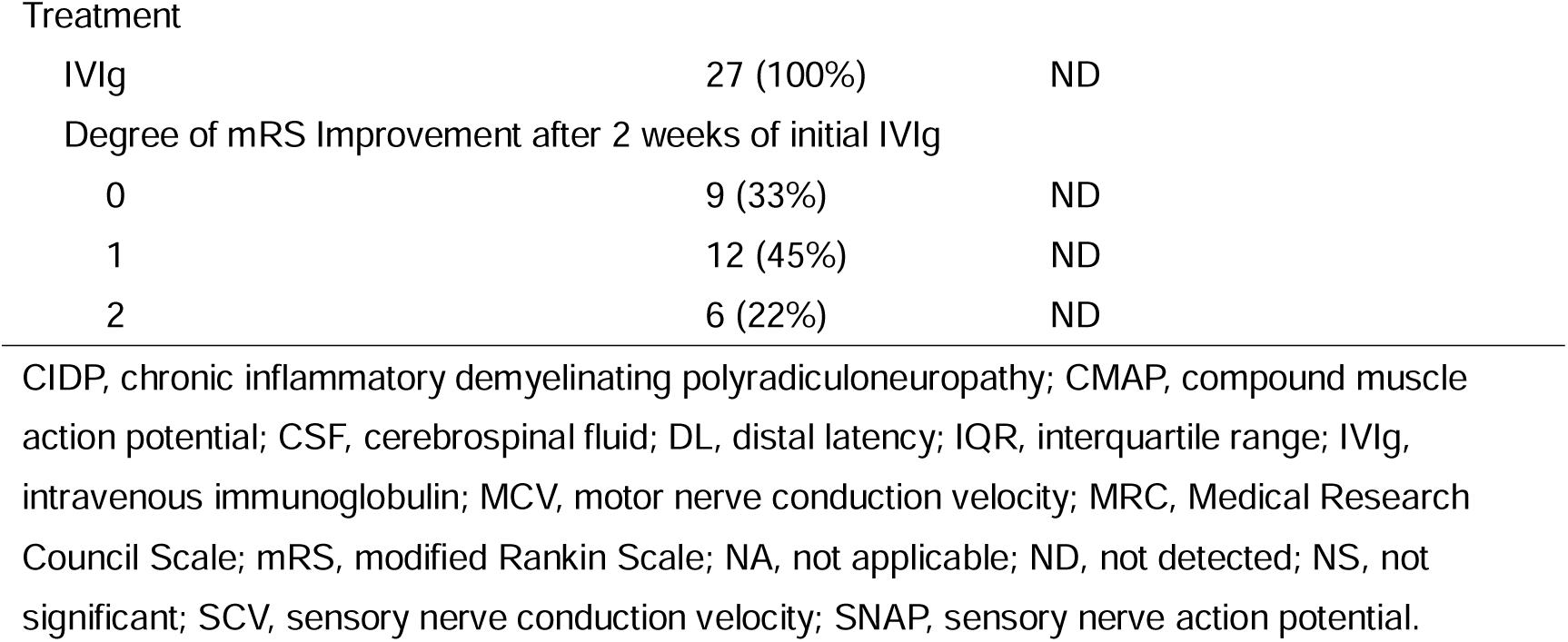
Characteristics of patients with CIDP and healthy controls.

|  | Typical CIDP<br>n=27 | Control<br>n=20 | P values |
| --- | --- | --- | --- |
| Age at sampling, y [median(IQR)] | 56 (36–69) | 66 (29–72) | NS |
| Sex, female | 13 (48%) | 9 (45%) | NS |
| Diabetes | 1 (4%) | 0 (0%) | NS |
| Disease duration, m [median(IQR)] | 6 (4–14) | NA |  |
| MRC scale | 44 (38–53) | ND |  |
| mRS | 3 (2–4) | 0 (0–0) | < 0.01 |
| Serum NfL levels, pg/mL [median(IQR)] | 13.2 (10.1–41.7) | 12.4 (5.9–17.3) | < 0.05 |
| CSF findings |  |  |  |
| Cell, /μl [median(IQR)] | 1 (1–2) | ND | |
| Protein, mg/dL [median(IQR)] | 73 (49–101) | ND |  |
| Motor conduction studies [median(IQR)] |  |  |  |
| Median nerve |  |  |  |
| DL, ms | 5.0 (3.9–6.1) | ND |  |
| CMAP amplitude, mV | 6.2 (3.7–7.7) | ND |  |
| MCV, m/s | 35.0 (29.0–45.0) | ND |  |
| Ulnar nerve |  |  |  |
| DL, ms | 3.4 (3.0–4.2) | ND |  |
| CMAP amplitude, mV | 5.7 (4.3–7.3) | ND |  |
| MCV, m/s | 41.0 (32.0–53.0) | ND |  |
| Tibial nerve |  |  |  |
| DL, ms | 5.3 (4.7–6.2) | ND |  |
| CMAP amplitude, mV | 5.2 (2.9–7.9) | ND |  |
| MCV, m/s | 37.0 (31.5–42.5) | ND |  |
| Peroneal nerve |  |  |  |
| DL, ms | 5.7 (4.9–6.8) | ND |  |
| CMAP amplitude, mV | 1.7 (0.4–2.3) | ND |  |
| MCV, m/s | 36.0 (32.3–43.0) | ND |  |
| Sensory conduction studies [median(IQR)] |  |  |  |
| Median nerve |  |  |  |
| SNAP amplitude, μV | 7.1 (1.8–11.4) | ND | |
| SCV, m/s | 44.4 (40.5–47.0) | ND |  |
| Ulnar nerve |  |  |  |
| SNAP amplitude, μV | 8.3 (0.4–14.4) | ND | |
| SCV, m/s | 41.0 (36.5–44.0) | ND |  |
| Sural nerve |  |  |  |
| SNAP amplitude, μV | 13.2 (2.9–21.8) | ND | |
| SCV, m/s | 44.1 (42.0–48.9) | ND |  |

|  |  |  |
| --- | --- | --- |
| Treatment |  |  |
| IVIg | 27 (100%) | ND |
| Degree of mRS Improvement after 2 weeks of initial IVIg |  |  |
| 0 | 9 (33%) | ND |
| 1 | 12 (45%) | ND |
| 2 | 6 (22%) | ND |

CIDP, chronic inflammatory demyelinating polyradiculoneuropathy; CMAP, compound muscle action potential; CSF, cerebrospinal fluid; DL, distal latency; IQR, interquartile range; IVIg, intravenous immunoglobulin; MCV, motor nerve conduction velocity; MRC, Medical Research Council Scale; mRS, modified Rankin Scale; NA, not applicable; ND, not detected; NS, not significant; SCV, sensory nerve conduction velocity; SNAP, sensory nerve action potential.

### Profiles of serum *N*- and *O*-type glycans by mass spectrometry analysis

In patients with CIDP and HC, 59 serum *N*-type and 18 serum *O*-type glycans were found (figure 2A, B). The former were classified into 5-high mannose, 39-sialylated, and 15-neutral types, whereas the latter were categorized into 11-sialylated and 7-neutral types. The sialylated glycans were divided into α2,3-, α2,6-, and α2,3- and α2,6-linked types according to their binding patterns. The most frequently observed *N*-glycan was the α2,6-linked sialylated type, Hex_2_HexNAc_2_NeuAc_2_ [α2,6/α2,6] +Man_3_GlcNAc_2_, and the most abundant *O*-glycan was α2,3-linked type, HexNAc_1_Hex_1_NeuAc_1_ [α2,3] (online supplementary tables e-1 and e-2).

**Figure 2.**
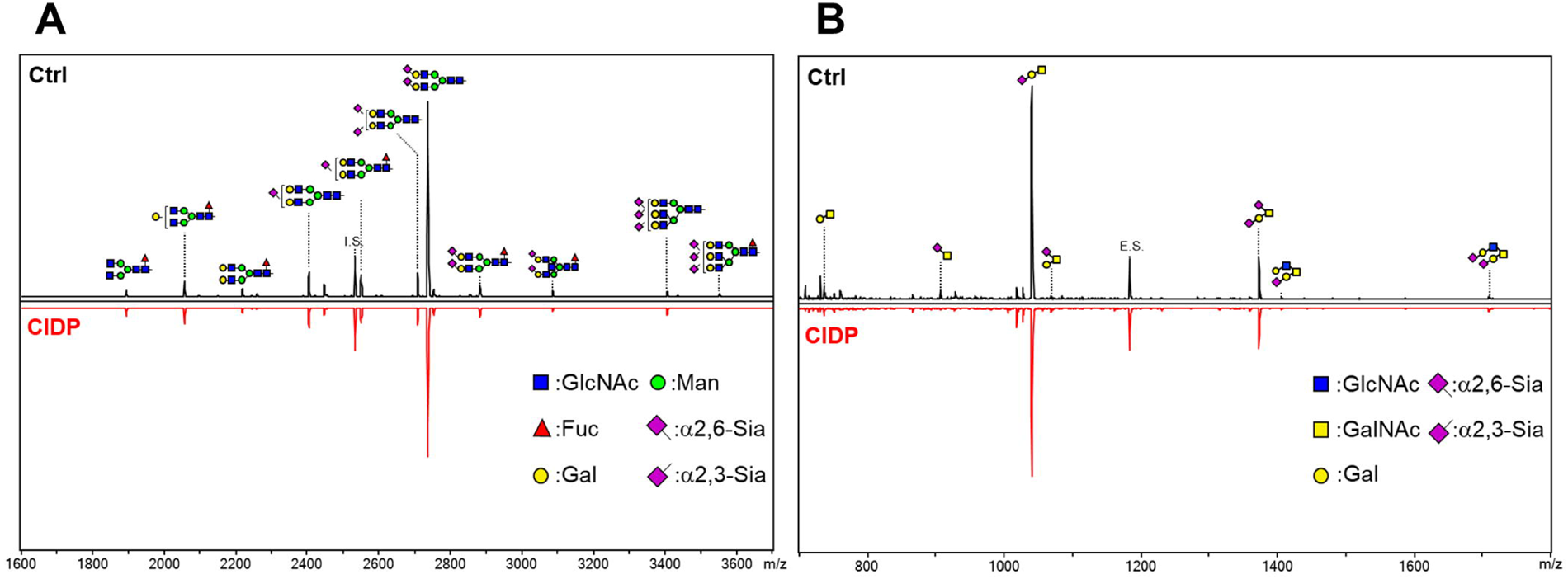
Representative serum *N*- and *O*-glycan profiles of patients with CIDP and controls. Representative matrix-assisted laser desorption-ionization time of flight mass spectrometry spectra of *N*-glycans (A) and *O*-glycans (B) in HC (Ctrl) and CIDP patient sera. Each peak was assigned a representative glycan structure.

### Comparison of the serum *N*- and *O*-glycan levels between the HC and CIDP patients

The serum total *N*-glycans levels of patients with CIDP were significantly lower than in the HC (CIDP, median 973.3 [IQR 836.2–1131.3] pmol/µL; HC, 1125.0 [1005.0–1236.2] pmol/µL; *p* < 0.05; figure 3A, B). In the subgroup analysis, the sialylated *N*-glycans levels were significantly lower in the patients with CIDP than in the HC (CIDP, 898.0 [752.2–1037.2] pmol/µL; HC, 1064.4 [942.7–1189.8] pmol/µL; p < 0.01), although the high mannose or neutral types did not differ significantly (figure 3C–E). The levels of α2,3-, α2,6-, and α2,3- and α2,6-linked sialylated *N*-glycans were all significantly lower in the patients with CIDP than in the HC (figure 3F–H). The ROC curve indicated that the serum total *N*-glycans, specifically sialylated types, were used to distinguish the patients with CIDP from the HC, with the respective AUC values of 0.704 (95% confidence interval [CI], 0.551–0.856, p < 0.05) and 0.735 (95% CI, 0.589–0.881, *p* < 0.01) (figure 3I).

**Figure 3.**
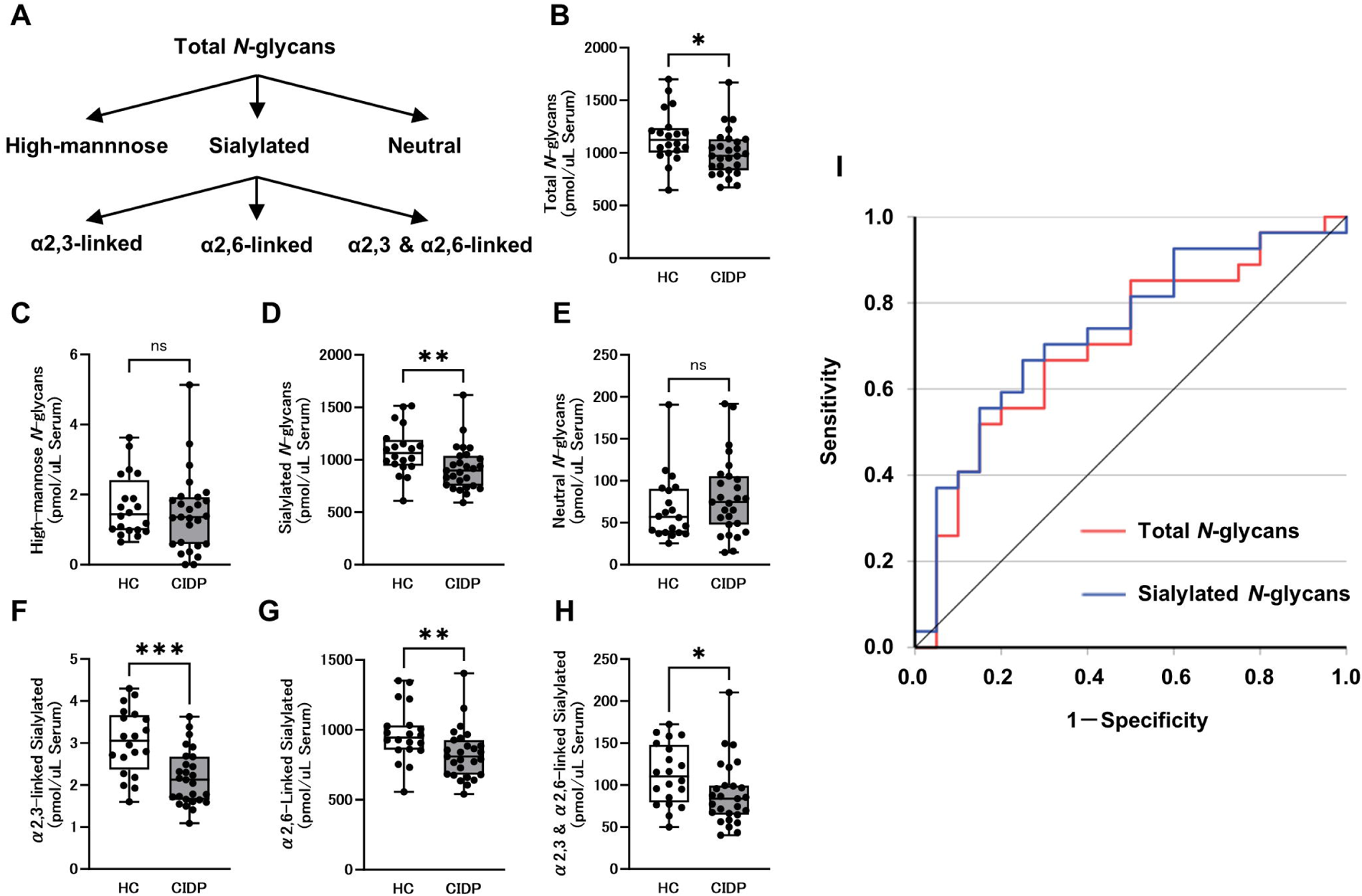
Serum *N*-glycans levels in patients with CIDP. **A**. Schema of *N*-glycan classification. **B.** Serum total *N*-glycans levels in the HC and patients with CIDP. **C–E.** Serum levels of the *N*-glycan subclass of high mannose (C), sialylated (D), and neutral (E) types. **F–H.** Serum levels of the sialylated *N*-glycan subclass of α 2, 3-(F), α2,6-(G), and α2,3- and α2,6-linked sialylated (H) glycans. **I.** Receiver operating characteristic (ROC) analysis discriminating CIDP patients from HC showed an area under the curve (AUC) for serum total *N*-glycans and sialylated *N*-glycans levels being 0.704 and 0.735, respectively. Horizontal lines in the boxplots indicate the median. The top and bottom edges of each box indicate the IQR. The I-bar indicates 1.5 times the IQR. Statistical analysis was performed using the Mann–Whitney U test. \**p* < 0.05, \*\**p* < 0.01, \*\*\**p* < 0.001.

The serum total *O*-glycans levels did not differ significantly between the CIDP and HC patients (figure 4A, B). Similarly, none of the subgroups, including the sialylated glycans, revealed a similar trend (figure 4C–G). Furthermore, the ROC curves did not contribute to the discrimination between the CIDP patients and HC with AUC values of 0.569 (95% CI, 0.402–0.735, *p* = 0.426) for total *O*-glycans and 0.581 (95% CI, 0.416–0.747, *p* = 0.344) for sialylated *O*-glycans (figure 4H).

**Figure 4.**
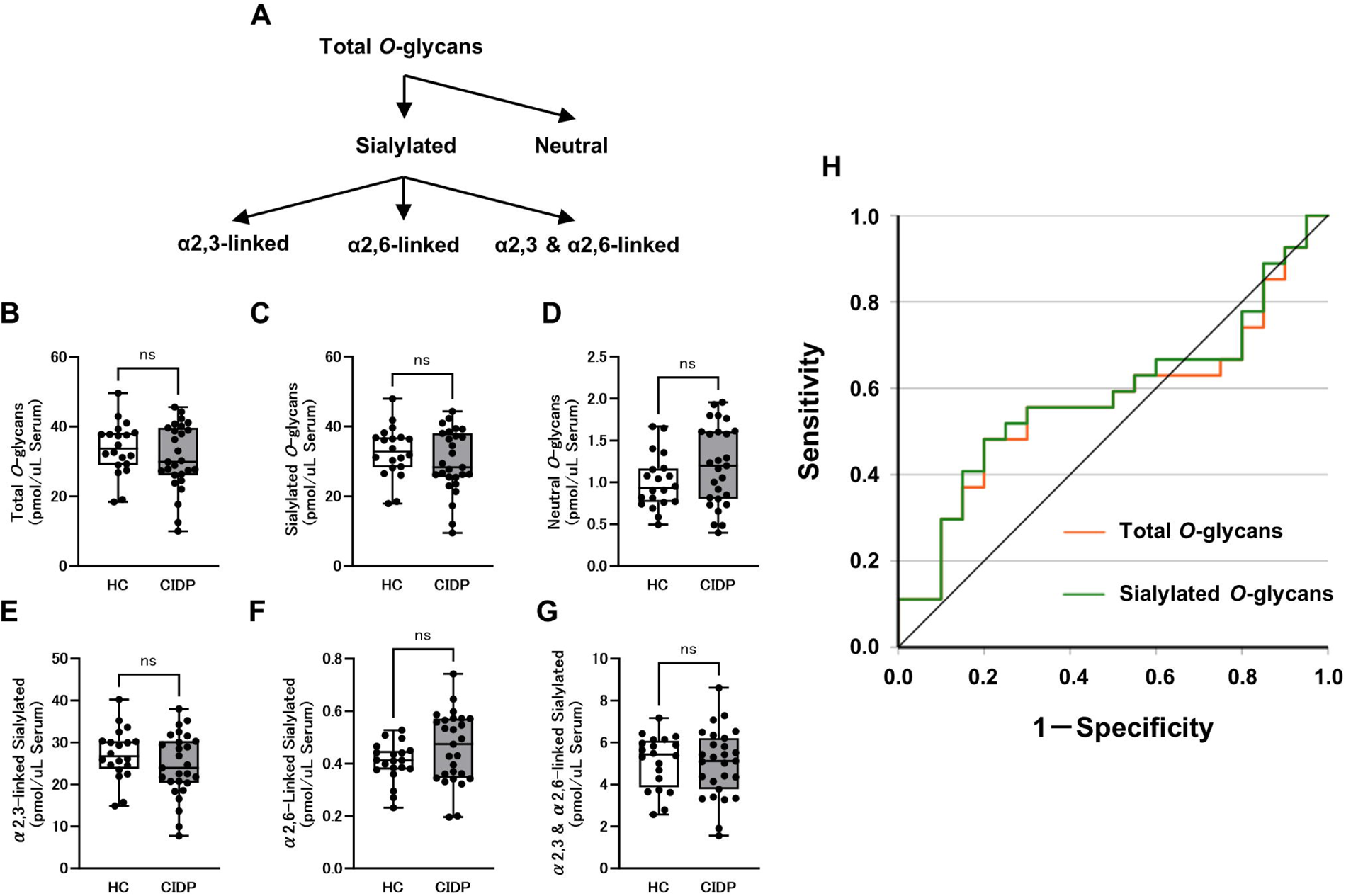
Serum *O*-glycans levels in patients with CIDP. **A**. Schema of *O*-glycan classification. **B.** Serum total *O*-glycans levels in the HC and CIDP patients. **C, D**, Serum levels of the *O*-glycan subclass of sialylated (C) and neutral (D) types. **E–G.** Serum levels of the sialylated *O*-glycan subclass of α2,3-(E), α2,6-(F), and α2,3-and α2,6-linked sialylated (G) glycans. **H.** ROC analysis revealed that the serum total *O*-glycans levels did not contribute to the discrimination between CIDP patients and HC. The top and bottom edges of each box indicate the IQR. The I-bar indicates 1.5 times the IQR. Statistical analysis was performed using the Mann–Whitney U test.

### Association between the serum *N*- and *O*-glycan levels and initial treatment response

To determine whether the serum glycan levels can predict susceptibility to treatment, we analyzed the correlation between the glycan levels and improvement in mRS scores as compared with the values at baseline and during the 2-week period after initial IVIg therapy. The significant difference in serum total *N*-glycans levels was observed among the three groups with mRS improvement scores of 0, 1, and 2 (801.1 [720.0–969.8], 1032.9 [878.9–1114.0], and 1163.2 [967.7–1405.4], respectively, *p* < 0.05) (figure 5A). After adjusting for multiple comparisons using Bonferroni correction, a significant difference was observed between Groups 0 and 2 (*p* < 0.05), indicating that lower *N*-glycan levels were associated with therapeutic resistance to initial IVIg therapy. In the subgroup analysis, the significant difference in the high mannose-type and sialylated glycans levels was observed among the three groups (figure 5B, C). There were no significant differences among groups in neutral types (figure 5D). In the sialylated glycans, the α2,6-linked type revealed a significant difference, but not in the α2,3- or α2,3- and α2,6-linked types (figure 5E–G). The ROC curve indicated that the serum total *N*-glycans, specifically the sialylated types, were distinguishable in the non-responder and responder groups, with respective AUC values of 0.827 (95% CI, 0.622–1.000, *p* < 0.01) and 0.840 (95% CI, 0.640–1.000, *p* < 0.01) (figure 5H).

**Figure 5.**
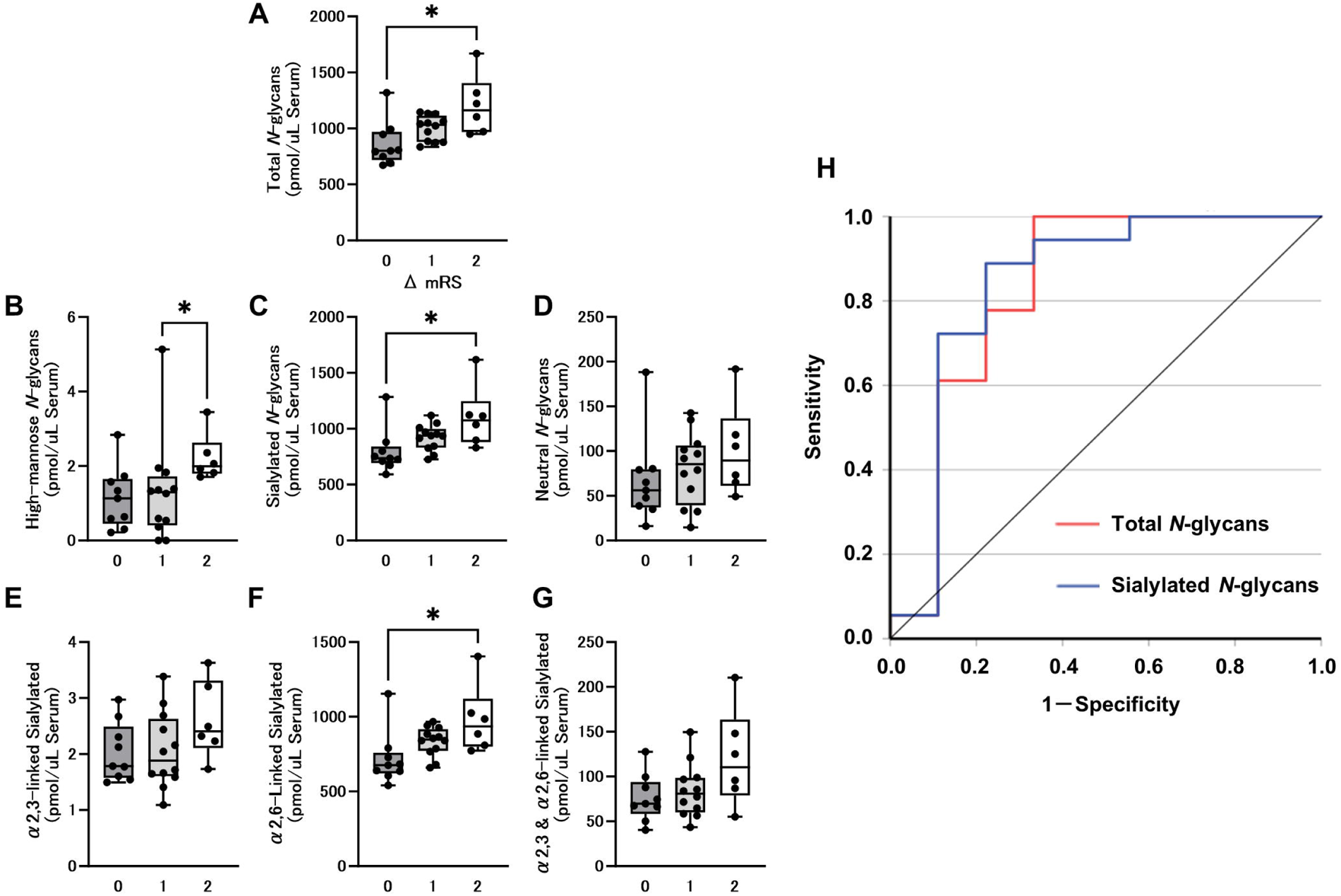
Association of serum *N*-glycans levels with response to initial IVIg treatment. A. Association between the serum total *N*-glycans levels and mRS improvement score from baseline after 2 weeks of initial IVIg treatment in patients with CIDP. B–D. Serum levels of *N*-glycan subgroup of high mannose (B), sialylated (C), and neutral (D) types. E, F. Serum levels of the sialylated *N*-glycan subgroup of α2,3-(E), α2,6-(F), and α2,3- and α2,6-linked sialylated (G) glycans. H. ROC analysis differentiating the responder group from the non-responder group showed that the AUC for the serum total *N*-glycans and sialylated *N*-glycans levels were 0.827 and 0.840, respectively. The top and bottom edges of each box indicate the IQR. The I-bar indicates 1.5 times the IQR. \**p* < 0.05, multiple comparisons using Bonferroni correction. ΔmRS, degree of mRS improvement after 2 weeks of initial IVIg.

There was a significant difference in serum total *O*-glycans levels among the three groups with mRS improvement scores of 0, 1, and 2 (23.8 [15.1–32.6], 30.1 [27.3–39.0], and 41.2 [36.7–44.6], respectively, *p* < 0.01) (figure 6A). Multiple comparisons revealed a significant difference between Groups 0 and 2. This indicated that lower glycan values, similar to *N*-glycans, were suggestive of therapeutic resistance. In the subgroup analysis, sialylated but not neutral glycans differed significantly among the three groups (figure 6B, C). A significant difference was observed in the α2,3-linked type, but not in the α2,6- and α2,3- and α2,6-linked types (figure 6D–F). The ROC curve indicated that the serum total *O*-glycans, especially sialylated *O*-glycans, could accurately discriminate between the non-responder and responder groups, showing respective AUC values of 0.846 (95% CI, 0.684–1.000, *p* < 0.01) and 0.846 (95% CI, 0.684–1.000, *p* < 0.01) (figure 6G).

**Figure 6.**
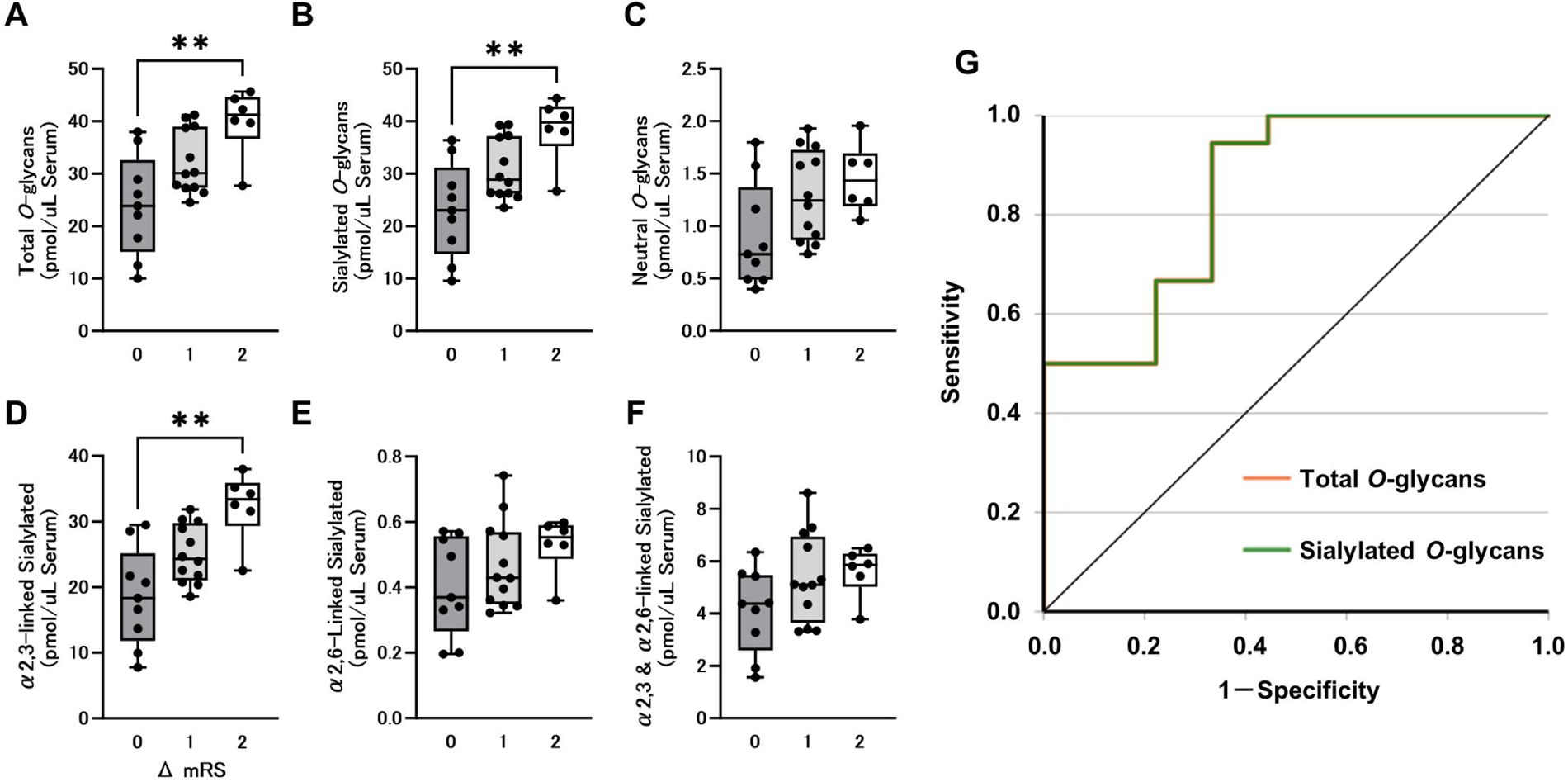
Association of serum *O*-glycans levels with response to initial IVIg treatment. A. Association between the serum total *O*-glycans levels and mRS improvement score from baseline after 2 weeks of initial IVIg treatment in patients with CIDP. B, C. Serum levels of *O*-glycan subgroup of sialylated (B) and neutral (C) types. D–F. Serum levels of sialylated *O*-glycan subgroup of α2,3- (D), α2,6- (E), and α2,3- and α2,6-linked sialylated (F) glycans. G. ROC analysis to discriminate the responder group from the non-responder group showed that the AUC for the serum total *O*-glycans and sialylated *O*-glycans levels were 0.846 and 0.846, respectively. The top and bottom edges of each box indicate the IQR. The I-bar indicates 1.5 times the IQR. \*\**p* < 0.01, multiple comparisons using Bonferroni correction. ΔmRS, degree of mRS improvement after 2 weeks of initial IVIg.

We analyzed the impact of other factors on treatment responsiveness (online supplementary figure e-1). Regarding the factors such as age and duration of illness, no significant differences were found among the three groups categorized by mRS improvement scores of 0, 1, and 2. Similarly, no significant differences in the levels of cerebrospinal fluid protein and CMAP amplitudes of the median, ulnar, peroneal, and tibial nerves were observed among the groups. Moreover, no significant difference in the serum NfL levels was observed among the groups.

### Identification of individual glycans that determine treatment responsiveness

To analyze the individual glycans responsible for therapeutic response, a discriminant model was generated using OPLS-DA. The non-responder and responder groups were distinctly separated based on the *N*- and *O*-glycans (figure 7A, B). The α2,6-linked sialylated *N*-glycans and α2,3-linked sialylated *O*-glycans exhibited high VIP scores (figure 7C, D). The ROC analysis was conducted for *N*-glycan, Hex_2_HexNAc_2_NeuAc_2_ [α2,6/α2,6] +Man_3_GlcNAc_2_, and *O*-glycan, Hex_2_HexNAc_2_. The AUC values from the ROC curves were 0.802 (95% CI, 0.595–1.000, *p* < 0.05) and 0.827 (95% CI, 0.673–0.982, *p* < 0.01), respectively, indicating high probability of discrimination.

**Figure 7.**
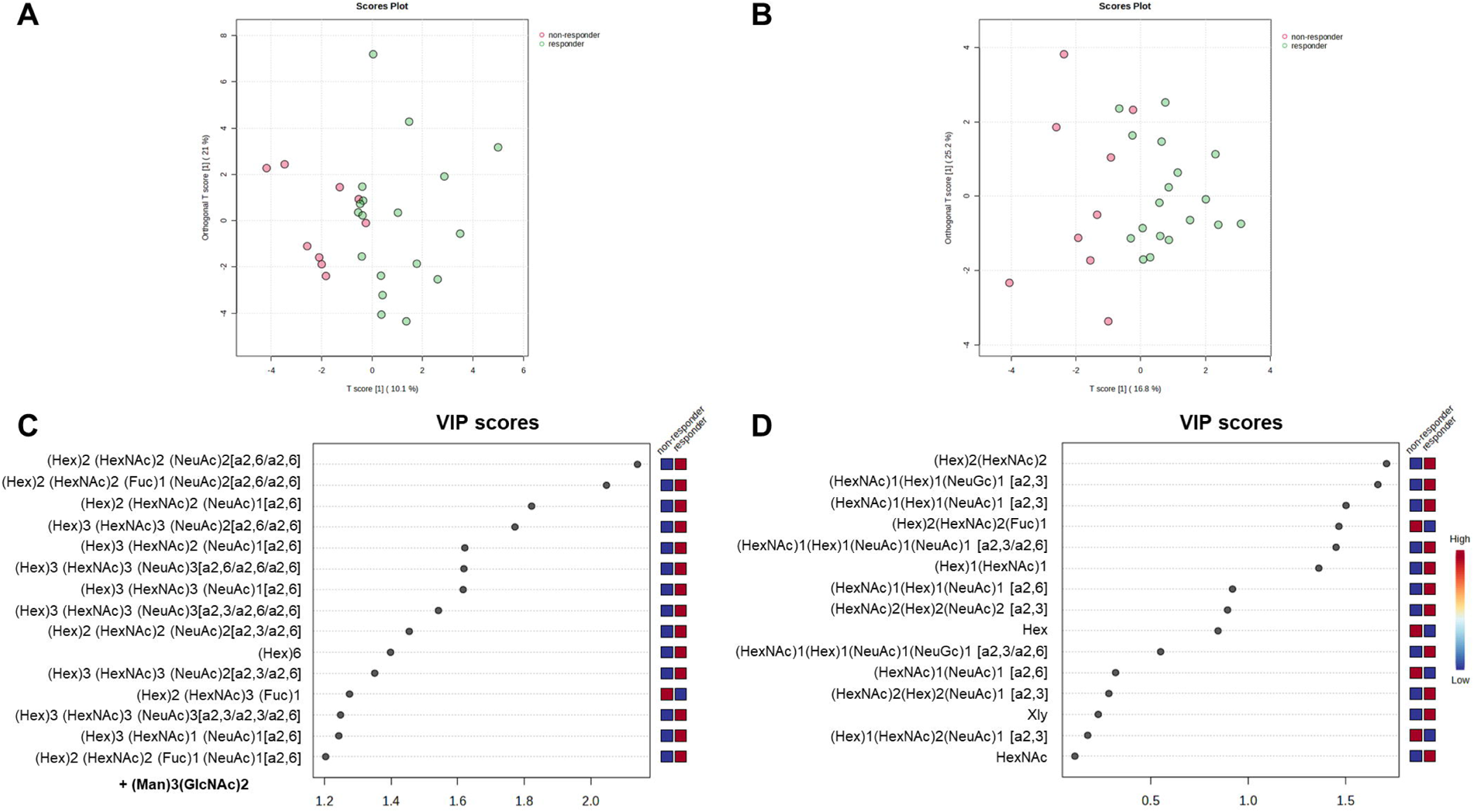
OPLS-DA model and VIP score of individual glycans. A, B. OPLS-DA score plot compares the glycomic patterns for the non-responder (red points) and responder groups (green points) in relation to *N*- (A) and *O*-glycans (B). C, D. VIP score plots of the top 15 glycans in *N*- (C) and *O*-glycans (D). VIP, Variable Importance in Projection.

### Correlation between the serum *N*- and *O*-glycans and electrophysiological indices

In the electrophysiological findings, a negative correlation was observed between the serum total *N*-glycans levels and distal latency of peroneal nerve (online supplementary table e-3). Furthermore, a positive correlation was observed between the serum total *O*-glycans levels and SCV of the sural nerve. In contrast, negative correlations were observed between the serum NfL levels and CMAP amplitude of the median, ulnar, or peroneal nerves. We observed a negative correlation between serum NfL levels and distal latency, and between serum NfL levels and MCV of the tibial and peroneal nerves. Moreover, a negative correlation was observed between the serum NfL levels and SNAP amplitude and SCV of the sural nerve.

No significant correlations between the *N*-glycans, *O*-glycans, and serum NfL levels were observed (*N*- and *O*-glycans, *rs* = 0.295 [*p* = 0.135]; *N*-glycans and serum NfL, *rs* = 0.153, [*p* = 0.447]; *O*-glycans and serum NfL, *rs* = −0.294 [*p* = 0.137]).

## DISCUSSION

In the present study, we performed a comprehensive analysis of the serum glycans of treatment-naïve patients with typical CIDP. We have discovered that the serum total *N*-glycans, specifically the sialylated types, were significantly lower in the patients with typical CIDP than in the HC. Furthermore, the lower levels of total *N*-glycans, particularly α2,6-sialylated *N*-glycans, and total *O*-glycans in this category of patients demonstrated a reduced responsiveness to initial IVIg treatment. This study reveals the potential of these glycans as novel biomarkers to determine the responsiveness to initial IVIg treatment.

In multiple sclerosis, multiple studies have reported that N-glycans play a crucial role in regulating oligodendrocytes and promoting myelin sheath formation and repair.^27–32^ In the serum of patients with progressive multiple sclerosis, a significant decrease in serum HexNAc, which is a stereoisomer of GlcNAc necessary for branching *N*-glycans, was observed. Lower levels of serum HexNAc have been reported to be positively correlated with disease severity.^28^ *N*-glycans were proven to suppress the responses of inflammatory type 1 and type 17 helper T-cells as well as activity of inflammatory B-cells in an inflammatory demyelination mouse model.^30^ Moreover, the oral supplementation of GlcNAc promoted remyelination and has been reported to exhibit neuroprotective effects for demyelinated axons in a mouse model.^31^ These studies suggest that a decrease in total serum *N*-glycan levels can serve as a potential biomarker of inflammation in the peripheral nervous system as well.

Regarding CIDP, the decreased levels of sialylated *N*-glycans within the IgG-Fc portion of the CIDP patients’ serum have been observed, revealing its association with disease severity.^20^ Furthermore, the presence of sialic acid in the *N*-glycans of IgG-Fc has been confirmed to compromise the effector function of inflammatory IgG, specifically by inhibiting complement-mediated cytotoxicity.^33^ The present study provides novel insights into the role of glycans in the development of CIDP: the decrease in sialylated *N*-glycans is indicative of an inflammatory pathophysiology in CIDP, affecting not only IgG-Fc but also the entire structure of glycoproteins.

Research on the association between therapeutic responsiveness to IVIg and serum glycosylation is limited. In the previous study, when IVIg was administered to the patients with CIDP, a significant elevation in serum sialylated IgG-Fc was observed showing a clinical improvement of the condition.^20^ Moreover, α2,6-linked sialylated *N*-glycans have been reported to be expressed more in M2 macrophages than in M1 macrophages and exhibit anti-inflammatory effects.^34^ In the present study, we discovered that the levels of α2,6-linked sialylated glycans were strongly associated with the therapeutic efficacy of IVIg.

In contrast to *N*-glycans, no significant difference was observed in the levels of *O*-glycans between the patients with typical CIDP and HC. Nevertheless, lower levels of *O*-glycans were linked with resistance to initial IVIg therapy. The dysregulation in the biosynthesis of *O*-glycans has been reported to play a crucial role in inflammation, infection, and carcinogenesis.^35, 36^ However, only a few studies on the role of *O*-type glycans in peripheral nerve disorders have been conducted, and several aspects remain unclear. Thus, further research in this area is required.

Serum NfL has been reported to be a useful biomarker reflecting axonal damage in CIDP.^18, 37^ In the present study, the serum NfL levels were discovered to be negatively correlated with CMAP amplitude in the nerves within the upper and lower limbs, possibly revealing axonal degeneration. An increase in serum NfL was confirmed to be associated with disease progression 1 year after serum sampling in patients with CIDP but has not been shown to reflect short-term treatment responses after IVIg administration.^38–40^ In our study, we also confirmed that serum NfL was not associated with treatment responsiveness to initial IVIg. Instead, the levels of serum glycans were associated with short-term initial response to IVIg treatment.

However, this study has several limitations. First, because of its exploratory nature and limited sample size, our findings should be interpreted with caution. The validation of our clinical observations can be strengthened by conducting further research with a larger cohort. Moreover, we did not assess other clinical outcomes, such as the Inflammatory Neuropathy Cause and Treatment disability score, Inflammatory Rasch-Built Overall Disability scale score, and grip strength. Furthermore, as this is a retrospective cohort research and post-treatment evaluations were missing, the long-term changes in glycan profiles remain uncertain. Finally, as the study focused exclusively on Japanese patients, glycans need to be measured in other ethnic groups to validate this marker.

In conclusion, our study identified low levels of serum total *N*-glycans, specifically the sialylated types, as potential glycan biomarkers for typical CIDP. Furthermore, they may predict the response to treatments with IVIg. However, a multicenter prospective longitudinal study is required to determine whether these glycans can serve as prognostic factors for the course of peripheral demyelinating diseases.

## Author contributions

Drafting/revising the manuscript for content: S.F., Y.F., J.F., C.S., and M.K. Study concept and design: S.F., Y.F., and M.K. Acquisition of samples, data, analysis and interpretation of data: S.F., Y.F., H.H., I.Y., J.F., M.H., K.K., C.S., K.H., Y.S., S.Y., H.K., and M.K.

## Acknowledgements

The authors would like to thank all the participants in this study and Enago, an editing brand of Crimson Interactive Inc. for proofreading a draft of this manuscript.

## Ethics approval

Ethics Review Committee of the Nagoya University Graduate School of Medicine, Nagoya, Japan.

## Data availability statement

All data produced in the present study are available upon reasonable request to the authors.

## Provenance and peer review

Not commissioned; externally peer reviewed.

