## Supplemental Data for "Serum Glycobiomarkers Defining Therapeutic Response to Intravenous Immunoglobulin in Chronic Inflammatory Demyelinating Polyneuropathy"

**Contents:**

Three Supplemental Tables (online supplementary table e-1, online supplementary table e-2, online supplementary table e-3)

One Supplemental Figure (online supplementary figure e-1)

***Corresponding author**: Masahisa Katsuno, Department of Neurology, Nagoya University Graduate School of Medicine,65　Tsurumai-Cho, Showa-ku, Nagoya, Aichi, 466-8550, Japan. Tel: +81-52-744-2389; Fax: +81-52-744-2384,

**Online supplementary table e-1. *N*-glycan Profile in serum of healthy controls and CIDP patients**

| No. | m/z | Chemical composition | HC Mean  ± SD  pmol/ μL | CIDP Mean  ± SD  pmol/ μL | P value |
| --- | --- | --- | --- | --- | --- |
| **high mannose *N*-glycans** | | | | | |
| 1 | 1664.6536 | (Hex)2 + (Man)3(GlcNAc)2 | 0.3525 ± 0.2953 | 0.4368 ± 0.4813 | 0.763 |
| 2 | 1826.7064 | (Hex)3 + (Man)3(GlcNAc)2 | 0.5634 ± 0.3455 | 0.4948 ± 0.3810 | 0.606 |
| 3 | 1988.7592 | (Hex)4 + (Man)3(GlcNAc)2 | 0.082 ± 0.0825 | 0.045 ± 0.0599 | 0.111 |
| 4 | 2150.812 | (Hex)5 + (Man)3(GlcNAc)2 | 0.1167 ± 0.1181 | 0.0987 ± 0.1386 | 0.411 |
| 5 | 2312.8648 | (Hex)6 + (Man)3(GlcNAc)2 | 0.5538 ± 0.2200 | 0.3851 ± 0.2454 | <0.05 |
| **neutral *N*-glycans** | | | | | |
| 6 | 1746.7068 | (HexNAc)2 + (Man)3(GlcNAc)2 | 0.0122 ± 0.0312 | 0.0047 ± 0.0171 | 0.387 |
| 7 | 1867.733 | (Hex)2 (HexNAc)1 + (Man)3(GlcNAc)2 | 0.0051 ± 0.0229 | 0.0027 ± 0.0142 | 0.806 |
| 8 | 1892.7647 | (HexNAc)2 (Fuc)1 + (Man)3(GlcNAc)2 | 13.3015 ± 15.4073 | 22.6832 ± 21.9390 | <0.05 |
| 9 | 1908.7596 | (Hex)1 (HexNAc)2 + (Man)3(GlcNAc)2 | 0.1554 ± 0.1818 | 0.1030 ± 0.1223 | 0.464 |
| 10 | 1949.7862 | (HexNAc)3 + (Man)3(GlcNAc)2 | 0.0338 ± 0.122 | 0.0392 ± 0.0811 | 0.321 |
| 11 | 2029.7858 | (Hex)3 (HexNAc)1 + (Man)3(GlcNAc)2 | 0.0168 ± 0.0421 | 0.0094 ± 0.0276 | 0.603 |
| 12 | 2054.8175 | (Hex)1 (HexNAc)2 (Fuc)1 + (Man)3(GlcNAc)2 | 30.6019 ± 15.9878 | 36.5518 ± 18.9787 | 0.322 |
| 13 | 2070.8124 | (Hex)2 (HexNAc)2 + (Man)3(GlcNAc)2 | 0.5108 ± 0.2852 | 0.3511 ± 0.2185 | 0.121 |
| 14 | 2095.8441 | (HexNAc)3 (Fuc)1 + (Man)3(GlcNAc)2 | 1.4634 ± 1.714 | 2.0658 ± 2.2047 | 0.138 |
| 15 | 2111.839 | (Hex)1 (HexNAc)3 + (Man)3(GlcNAc)2 | 0.0334 ± 0.0721 | 0.0802 ± 0.1574 | 0.313 |
| 16 | 2216.8703 | (Hex)2 (HexNAc)2 (Fuc)1 + (Man)3(GlcNAc)2 | 14.9131 ± 8.593 | 12.8980 ± 7.1733 | 0.478 |
| 17 | 2257.8969 | (Hex)1 (HexNAc)3 (Fuc)1 + (Man)3(GlcNAc)2 | 3.7824 ± 2.9545 | 4.4245 ± 3.2181 | 0.282 |
| 18 | 2273.8918 | (Hex)2(HexNAc)3+(Man)3(GlcNAc)2 | 0.0227 ± 0.0567 | 0.0113 ± 0.0413 | 0.410 |
| 19 | 2419.9497 | (Hex)2 (HexNAc)3 (Fuc)1 + (Man)3(GlcNAc)2 | 2.0514 ± 1.1238 | 1.4388 ± 0.7359 | <0.05 |
| 20 | 2582.0025 | (Hex)3 (HexNAc)3 (Fuc)1 + (Man)3(GlcNAc)2 | 0.0087 ± 0.0389 | 0.0000 ± 0.0000 | 0.245 |
| **sialylated *N*-glycans** | | | | | |
| 21 | 2374.94366 | (Hex)2 (HexNAc)2 (NeuAc)1[a2,3] + (Man)3(GlcNAc)2 | 0.8252 ± 0.3683 | 0.6237 ± 0.2533 | <0.05 |
| 22 | 2505.00666 | (Hex)1 (HexNAc)2 (Fuc)2 (NeuAc)1[a2,3] + (Man)3(GlcNAc)2 | 0.0000 ± 0.0000 | 0.0169 ± 0.0441 | 0.075 |
| 23 | 2521.00156 | (Hex)2 (HexNAc)2 (Fuc)1 (NeuAc)1[a2,3] + (Man)3(GlcNAc)2 | 1.0061 ± 0.2782 | 0.7570 ± 0.2099 | <0.01 |
| 24 | 2667.05946 | (Hex)2 (HexNAc)2 (Fuc)2 (NeuAc)1[a2,3] + (Man)3(GlcNAc)2 | 0.0000 ± 0.0000 | 0.0057 ± 0.0295 | 0.389 |
| 25 | 2679.07492 | (Hex)2 (HexNAc)2 (NeuAc)2[a2,3/a2,3] + (Man)3(GlcNAc)2 | 0.3476 ± 0.0853 | 0.1694 ± 0.1619 | <0.001 |
| 26 | 2724.08096 | (Hex)2 (HexNAc)3 (Fuc)1 (NeuAc)1[a2,3] + (Man)3(GlcNAc)2 | 0.0505 ± 0.0914 | 0.0120 ± 0.0439 | 0.086 |
| 27 | 2825.13282 | (Hex)2 (HexNAc)2 (Fuc)1 (NeuAc)2[a2,3/a2,3] + (Man)3(GlcNAc)2 | 0.7992 ± 0.1877 | 0.5869 ± 0.1915 | <0.001 |
| 28 | 1875.78996 | (HexNAc)1 (NeuAc)1[a2,6] + (Man)3(GlcNAc)2 | 0.0159 ± 0.0406 | 0.0038 ± 0.0145 | 0.345 |
| 29 | 2037.84276 | (Hex)1 (HexNAc)1 (NeuAc)1[a2,6] + (Man)3(GlcNAc)2 | 0.2807 ± 0.1829 | 0.1983 ± 0.1652 | 0.116 |
| 30 | 2183.90066 | (Hex)1 (HexNAc)1 (Fuc)1 (NeuAc)1[a2,6]+ (Man)3(GlcNAc)2 | 0.0046 ± 0.0206 | 0.0078 ± 0.0282 | 0.703 |
| 31 | 2199.89556 | (Hex)2 (HexNAc)1 (NeuAc)1[a2,6] + (Man)3(GlcNAc)2 | 0.5458 ± 0.2668 | 0.5266 ± 0.3044 | 0.667 |
| 32 | 2240.92216 | (Hex)1 (HexNAc)2 (NeuAc)1[a2,6] + (Man)3(GlcNAc)2 | 0.7768 ± 0.6772 | 0.7594 ± 0.6438 | 0.897 |
| 33 | 2361.94836 | (Hex)3 (HexNAc)1 (NeuAc)1[a2,6] + (Man)3(GlcNAc)2 | 0.3527 ± 0.1635 | 0.2463 ± 0.1817 | <0.05 |
| 34 | 2386.98006 | (Hex)1 (HexNAc)2 (Fuc)1 (NeuAc)1[a2,6] + (Man)3(GlcNAc)2 | 1.0207 ± 0.4618 | 1.1189 ± 0.6418 | 0.897 |
| 35 | 2402.97496 | (Hex)2 (HexNAc)2 (NeuAc)1[a2,6] + (Man)3(GlcNAc)2 | 94.7950 ± 39.4059 | 78.7563 ± 24.9703 | 0.175 |
| 36 | 2549.03286 | (Hex)2 (HexNAc)2 (Fuc)1 (NeuAc)1[a2,6] + (Man)3(GlcNAc)2 | 39.5540 ± 15.6253 | 32.5009 ± 12.3687 | 0.175 |
| 37 | 2565.02776 | (Hex)3 (HexNAc)2 (NeuAc)1[a2,6] + (Man)3(GlcNAc)2 | 0.3440 ± 0.3028 | 0.1603 ± 0.2165 | <0.05 |
| 38 | 2590.05946 | (Hex)1 (HexNAc)3 (Fuc)1 (NeuAc)1[a2,6] + (Man)3(GlcNAc)2 | 0.3705 ± 0.3625 | 0.5628 ± 0.7233 | 0.522 |
| 39 | 2606.05436 | (Hex)2 (HexNAc)3 (NeuAc)1[a2,6] + (Man)3(GlcNAc)2 | 1.3613 ± 1.1682 | 1.2988 ± 1.3874 | 0.505 |
| 40 | 2695.09076 | (Hex)2 (HexNAc)2 (Fuc)2 (NeuAc)1[a2,6] + (Man)3(GlcNAc)2 | 0.0000 ± 0.0000 | 0.07170 ± 0.3726 | 0.389 |
| 41 | 2735.13752 | (Hex)2 (HexNAc)2 (NeuAc)2[a2,6/a2,6] + (Man)3(GlcNAc)2 | 772.6195 ± 157.6319 | 662.1118 ± 165.3016 | <0.05 |
| 42 | 2752.11226 | (Hex)2 (HexNAc)3 (Fuc)1 (NeuAc)1[a2,6] + (Man)3(GlcNAc)2 | 23.7021 ± 14.1587 | 17.0229 ± 9.9946 | 0.074 |
| 43 | 2768.10716 | (Hex)3 (HexNAc)3 (NeuAc)1[a2,6] + (Man)3(GlcNAc)2 | 1.9139 ± 0.9426 | 1.0670 ± 0.8065 | <0.01 |
| 44 | 2881.19542 | (Hex)2 (HexNAc)2 (Fuc)1 (NeuAc)2[a2,6/a2,6] + (Man)3(GlcNAc)2 | 20.9835 ± 10.0858 | 19.4358 ± 9.1872 | 0.505 |
| 45 | 2914.16506 | (Hex)3 (HexNAc)3 (Fuc)1 (NeuAc)1[a2,6] + (Man)3(GlcNAc)2 | 0.1600 ± 0.1184 | 0.0888 ± 0.0890 | <0.05 |
| 46 | 2938.21692 | (Hex)2 (HexNAc)3 (NeuAc)2[a2,6] + (Man)3(GlcNAc)2 | 0.0634 ± 0.1044 | 0.0249 ± 0.0616 | 0.167 |
| 47 | 3084.27482 | (Hex)2 (HexNAc)3 (Fuc)1 (NeuAc)2[a2,6/a2,6] + (Man)3(GlcNAc)2 | 7.8254 ± 7.1791 | 9.684 ± 12.4826 | 0.830 |
| 48 | 3100.26972 | (Hex)3 (HexNAc)3 (NeuAc)2[a2,6/a2,6] + (Man)3(GlcNAc)2 | 0.8090 ± 0.4930 | 0.5330 ± 0.4242 | <0.05 |
| 49 | 3246.32762 | (Hex)3 (HexNAc)3 (Fuc)1 (NeuAc)2[a2,6/a2,6] + (Man)3(GlcNAc)2 | 0.0312 ± 0.1063 | 0.0138 ± 0.0500 | 0.722 |
| 50 | 3432.43228 | (Hex)3 (HexNAc)3 (NeuAc)3[a2,6/a2,6/a2,6] + (Man)3(GlcNAc)2 | 3.0262 ± 2.5515 | 1.7734 ± 1.4863 | <0.05 |
| 51 | 2707.10622 | (Hex)2 (HexNAc)2 (NeuAc)2[a2,3/a2,6] + (Man)3(GlcNAc)2 | 66.9269 ± 18.8389 | 55.8431 ± 22.2790 | <0.05 |
| 52 | 2853.16412 | (Hex)2 (HexNAc)2 (Fuc)1 (NeuAc)2[a2,3/a2,6] + (Man)3(GlcNAc)2 | 2.1842 ± 1.0207 | 2.0977 ± 1.0568 | 0.846 |
| 53 | 3056.24352 | (Hex)2 (HexNAc)3 (Fuc)1 (NeuAc)2[a2,3/a2,6] + (Man)3(GlcNAc)2 | 0.0611 ± 0.1119 | 0.0358 ± 0.1150 | 0.237 |
| 54 | 3072.23842 | (Hex)3 (HexNAc)3 (NeuAc)2[a2,3/a2,6] + (Man)3(GlcNAc)2 | 1.2570 ± 0.7070 | 0.9688 ± 0.7841 | 0.085 |
| 55 | 3218.29632 | (Hex)3 (HexNAc)3 (Fuc)1 (NeuAc)2[a2,3/a2,6] + (Man)3(GlcNAc)2 | 0.0810 ± 0.1690 | 0.0464 ± 0.0887 | 0.662 |
| 56 | 3376.36968 | (Hex)3 (HexNAc)3 (NeuAc)3[a2,3/a2,3/a2,6] + (Man)3(GlcNAc)2 | 0.8991 ± 0.5469 | 0.6795 ± 0.6542 | 0.085 |
| 57 | 3404.40098 | (Hex)3 (HexNAc)3 (NeuAc)3[a2,3/a2,6/a2,6] + (Man)3(GlcNAc)2 | 34.7527 ± 19.0509 | 23.2955 ± 17.2103 | 0.058 |
| 58 | 3522.42758 | (Hex)3 (HexNAc)3 (Fuc)1 (NeuAc)3[a2,3/a2,3/a2,6] + (Man)3(GlcNAc)2 | 0.1016 ± 0.1820 | 0.0809 ± 0.1175 | 0.867 |
| 59 | 3550.45888 | (Hex)3 (HexNAc)3 (Fuc)1 (NeuAc)3[a2,3/a2,6/a2,6] + (Man)3(GlcNAc)2 | 6.4372 ± 9.9546 | 6.2858 ± 6.7304 | 0.846 |

P-values were calculated using the Mann-Whitney U test between the HC and CIDP groups (significance level α = 0.05). CIDP chronic inflammatory demyelinating polyradiculoneuropathy; HC, healty controls; m/z, mass-to-charge ratio.

**Online supplementary table e-2. *O*-glycan Profile in serum of healthy controls and CIDP patients**

| No. | m/z | Chemical composition | HC Mean  ± SD pmol/ μL | CIDP Mean  ± SD pmol/ μL | P value |
| --- | --- | --- | --- | --- | --- |
| **neutral *O*-glycans** | | | | | |
| 1 | 503.1909 | Xly | 0.0854 ± 0.0391 | 0.0778 ± 0.0436 | 0.547 |
| 2 | 533.2012 | Hex | 0.0278 ± 0.0446 | 0.0422 ± 0.0651 | 0.591 |
| 3 | 574.2280 | HexNAc | 0.0369 ± 0.0669 | 0.0507 ± 0.0757 | 0.595 |
| 4 | 736.2806 | (Hex)1(HexNAc)1 | 0.7747 ± 0.2619 | 0.9759 ± 0.4052 | 0.111 |
| 5 | 939.3600 | (Hex)1(HexNAc)2 | 0.0000 ± 0.0000 | 0.0000 ± 0.0000 | 1.000 |
| 6 | 1101.4126 | (Hex)2(HexNAc)2 | 0.0493 ± 0.0430 | 0.0400 ± 0.0442 | 0.509 |
| 7 | 1247.4705 | (Hex)2(HexNAc)2(Fuc)1 | 0.0279 ± 0.0333 | 0.0106 ± 0.0237 | <0.05 |
| **sialylated *O*-glycans** | | | | | |
| 8 | 906.3863 | (HexNAc)1(6NeuAc)1[a2,6] | 0.0161 ± 0.0309 | 0.0167 ± 0.0413 | 0.562 |
| 9 | 1040.4076 | (HexNAc)1(Hex)1(3NeuAc)1[a2,3] | 26.3276 ± 5.9795 | 23.4743 ± 7.4477 | 0.182 |
| 10 | 1056.4025 | (HexNAc)1(Hex)1(3NeuGc)1[a2,3] | 0.1930 ± 0.0542 | 0.2121 ± 0.0901 | 0.333 |
| 11 | 1068.4389 | (HexNAc)1(Hex)1(6NeuAc)1[a2,6] | 0.3883 ± 0.0694 | 0.4433 ± 0.1240 | 0.102 |
| 12 | 1372.5659 | (HexNAc)1(Hex)1(6NeuAc)1(3NeuAc)1　[a2,3/a2,6] | 5.0641 ± 1.2535 | 4.9766 ± 1.6104 | 0.813 |
| 13 | 1388.5608 | (HexNAc)1(Hex)1(6NeuAc)1(3NeuGc)1　[a2,3/a2,6] | 0.0366 ± 0.0446 | 0.0270 ± 0.0446 | 0.345 |
| 14 | 1243.4870 | (Hex)1(HexNAc)2(3NeuAc)1[a2,3] | 0.0398 ± 0.0329 | 0.0246 ± 0.0391 | 0.107 |
| 15 | 1405.5396 | (HexNAc)2(Hex)2(3NeuAc)1[a2,3] | 0.1836 ± 0.0447 | 0.2517 ± 0.0965 | <0.01 |
| 16 | 1551.5975 | (HexNAc)2(Hex)2(3NeuAc)1(Fuc)1[a2,3] | 0.0288 ± 0.0294 | 0.0135 ± 0.0271 | <0.05 |
| 17 | 1709.6666 | (HexNAc)2(Hex)2(3NeuAc)2[a2,3] | 0.4163 ± 0.1077 | 0.5088 ± 0.1669 | <0.05 |
| 18 | 1855.7245 | (HexNAc)2(Hex)2(3NeuAc)2(Fuc)1[a2,3] | 0.0277 ± 0.0390 | 0.0290 ± 0.0508 | 0.726 |

P-values were calculated using the Mann-Whitney U test between the HC and CIDP groups (significance level α = 0.05). m/z, mass-to-charge ratio.

**Online supplementary table e-3. Correlation of nerve conduction studies with serum *N*- or *O*-glycan and neurofilament levels**

|  |  | *N*-glycans | |  | *O*-glycans | |  | NfL | |
| --- | --- | --- | --- | --- | --- | --- | --- | --- | --- |
|  |  | *rs* | *p* |  | *rs* | *p* |  | *rs* | *p* |
| DL (ms) | Median nerve | ‐0.315 | 0.110 |  | ‐0.167 | 0.405 |  | 0.164 | 0.413 |
|  | Ulnar nerve | ‐0.190 | 0.341 |  | 0.017 | 0.935 |  | 0.271 | 0.172 |
|  | Tibial nerve | ‐0.158 | 0.450 |  | 0.013 | 0.949 |  | ‐0.501 | 0.011 |
|  | Peroneal nerve | ‐0.524 | 0.010 |  | ‐0.160 | 0.466 |  | ‐0.411 | 0.052 |
| CMAP amp. (mV) | Median nerve | 0.087 | 0.665 |  | 0.139 | 0.489 |  | ‐0.455 | 0.017 |
|  | Ulnar nerve | 0.122 | 0.544 |  | 0.067 | 0.739 |  | ‐0.501 | 0.008 |
|  | Tibial nerve | 0.109 | 0.587 |  | 0.000 | 0.999 |  | ‐0.361 | 0.064 |
|  | Peroneal nerve | 0.162 | 0.421 |  | 0.212 | 0.288 |  | ‐0.507 | 0.007 |
| MCV (m/s) | Median nerve | 0.172 | 0.391 |  | ‐0.102 | 0.612 |  | ‐0.024 | 0.904 |
|  | Ulnar nerve | 0.126 | 0.531 |  | ‐0.218 | 0.274 |  | 0.120 | 0.552 |
|  | Tibial nerve | 0.108 | 0.608 |  | 0.214 | 0.304 |  | ‐0.515 | 0.008 |
|  | Peroneal nerve | ‐0.029 | 0.894 |  | 0.220 | 0.301 |  | ‐0.541 | 0.006 |
| SNAP amp. (μV) | Median nerve | 0.099 | 0.622 |  | ‐0.096 | 0.632 |  | ‐0.184 | 0.359 |
|  | Ulnar nerve | 0.097 | 0.630 |  | ‐0.257 | 0.195 |  | ‐0.199 | 0.319 |
|  | Sural nerve | ‐0.259 | 0.192 |  | 0.133 | 0.509 |  | ‐0.655 | <0.001 |
| SCV (m/s) | Median nerve | 0.336 | 0.137 |  | 0.026 | 0.911 |  | ‐0.059 | 0.801 |
|  | Ulnar nerve | 0.147 | 0.525 |  | ‐0.307 | 0.175 |  | 0.026 | 0.911 |
|  | Sural nerve | 0.186 | 0.407 |  | 0.530 | 0.011 |  | ‐0.494 | 0.019 |

CMAP amp, compound muscle action potential amplitude; DL, distal latency; MCV, motor nerve conduction velocity; NfL, neurofilament light chain; SCV, sensory nerve conduction velocity; SNAP amp, sensory nerve action potential amplitude.

**Online supplementary figure e-1. Association of factors other than glycans with the initial response to treatment with IVIg.**

B

A

C

D

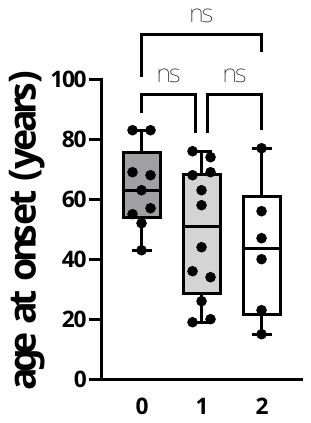

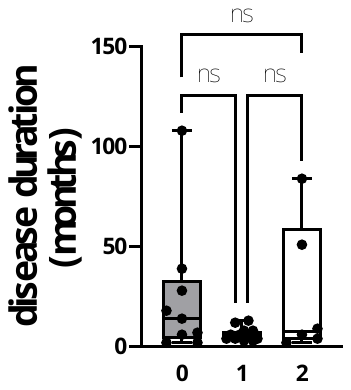

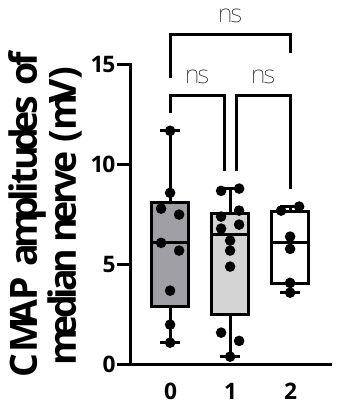

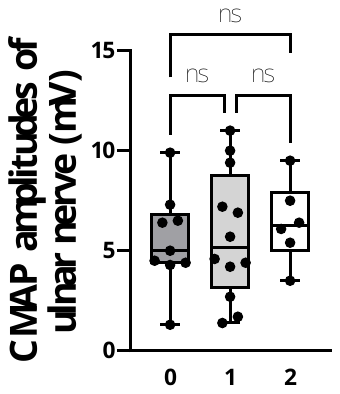

Δ mRS

H

E

F

G

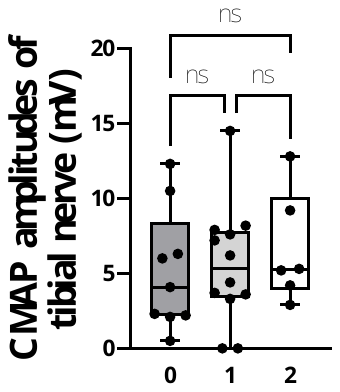

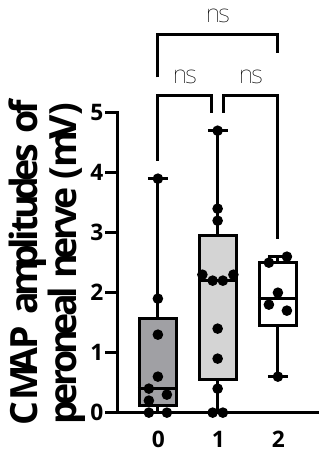

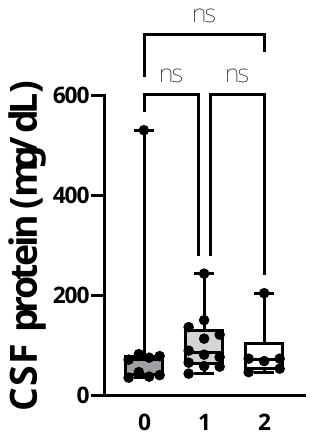

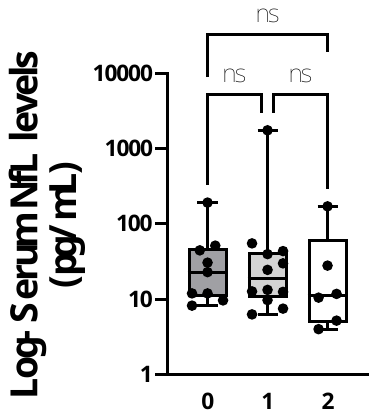

**A­­­­–H.** There were no significant differences in age (A), duration of disease (B), CMAP amplitudes of each nerve (C–F), CSF protein (G), nor serum NfL levels (H) among the three groups with mRS improvements of 0, 1 and 2. Statistical analysis was performed using the Kruskal-Wallis test. Horizontal lines in the boxplots indicate the median. The top and bottom edges of each box indicate the IQR. The I-bar indicates 1.5 times the IQR. ns, not significant; ΔmRS, degree of mRS improvement after 2 weeks of initial IVIg.
